# High prevalence of Human Papillomavirus (HPV) type 66 in low-grade cervical lesions of Mexican women

**DOI:** 10.1101/2020.01.07.20016857

**Authors:** Karina Juárez-González, Vladimir Paredes-Cervantes, Silvia Gordillo-Rodríguez, Saúl González-Guzmán, Xochilt Moncayo-Valencia, Rocío Méndez-Martínez, Alejandro García-Carrancá, José Darío Martínez-Ezquerro, Rodolfo Rivas-Ruiz, Patricia Sánchez-Suárez, Paola Álvarez-Sandoval, Patricia Padilla-Arrieta, Martha Martínez-Salazar, Salvador Vázquez-Vega

**Author notes:** Equal contribution (Karina Juárez-González, Vladimir Paredes-Cervantes). Corresponding authors: Salvador Vázquez-Vega, Unidad de Investigación Epidemiológica y en Servicios de Salud, CMN Siglo XXI, IMSS. Av. Cuauhtémoc, 330. Col. Doctores Alcaldía Cuauhtémoc. Ciudad de México. CP. 06720. México.: (52., Martha Martínez-Salazar, Unidad de Investigación Médica en Inmunoquímica. UMAE Especialidades CMN Siglo XXI, IMSS. Av. Cuauhtémoc, 330. Col. Doctores Alcaldía Cuauhtémoc. Ciudad de México. CP. 06720. México.: (52.

## Abstract

**Background:** HPV-16 infections constitute the highest risk for developing uterine cervix cancer. However, the role of other high-risk types is still controversial.

**Objective:** To analyze HR-HPV prevalence and its possible associations between HPV and risk factors related to cervical lesions among Mexican women.

**Methods:** Cross sectional study using 362 cervical samples collected between 2016 and 2017. Fourteen HR-HPV types (16, 18, 31, 33, 35, 39, 45, 51, 52, 56, 58, 59, 66 and 68) were detected by highly sensitive PCR amplification followed by reverse hybridization. Bivariate and multivariate analyses were performed to investigate the association between HPV types and risk factors among lesions.

**Results:** Most samples were HR-HPV positive (83.43%). HPV-16 was the most prevalent infection among negative for intraepithelial lesions or malignancy (78.6%), high-grade squamous intraepithelial lesions (50%), and cervical cancer (58.2%). HPV-66 showed an unexpected high prevalence in atypical squamous cells of undetermined significance (50%), low-grade squamous intraepithelial (45.7%), and only found in 3.6% of cervical cancers. HPV-16 was significantly prevalent among women between 30-39 years, whereas types 66 and 52 were significantly associated when previously sexually transmitted disease had occurred (p< 0.05).

**Conclusions:** HPV-66 either in single or co-infection with other HR-HPV types (excluding 16 and 18) might be indicative of non-progressive cancer lesions. HPV-66 prevalence was unusually high in low-grade cervical lesions, predominantly in co-infection with HPV-51, and very low among cervical cancer. This should be addressed to interpret results obtained by methods that group type 66 with other HR-types.

## 1. Background

Cervical cancer (CC) is still a major health problem throughout the world, specifically in developing countries. In Mexico, it represents the second cause of death produced by gynecological malignancies [1], where CC has an incidence rate of 23.3 per 100,000 women and a mortality rate of 11.3 per 100,000 women [2]. It is well known that human papillomavirus (HPV) infections are the major sexually transmitted disease and that persistent infections with a set of high-risk HPV (HR-HPV) types constitute the principal etiological agent for developing CC [3]. Among these, HPV types 16, 18, 31, 33, 35, 39, 45, 51, 52, 56, 58, 59, 66, and 68 have been defined as HR-HPVs [4–6], based on their prevalence in cervical cancer, molecular characterization of viral type, and phylogenetic relationships between these viruses [4,7–9]. It must be taken into account that prevalence of HPV types, including HPV-66, is dependent on viral detection methods, age of participants, geographical distribution, sample type, among others [10].

HPV-66 was originally detected in a biopsy of a 38 year old patient with a stage I invasive squamous-cell carcinoma of the uterine cervix [11]. Several papers have reported the prevalence of HPV-66 between 4.6 - 36.5%, predominantly in premalignant lesions [10,12,13]. HPV-66 is an α-PV from species 6 and belonging to a third phylogenetic branch unrelated to types 16 and 18 [14]. Indeed, E6 and E7 proteins from type 66 have been shown to degrade p53 and RB cellular proteins, with lower efficiency than other HR-HPV types [15,16].

The aim of this study was to analyze the prevalence and possible associations between HR-HPVs in cervical lesions as well as the risk factors related with HR-HPV infections in Mexican women.

## 2. Methods

### 2.1. Study design

This transversal study was realized from March 2016 to February 2017 at the Gynecologic and Pediatric Hospital 3A and Oncology Hospital of Instituto Mexicano del Seguro Social (IMSS), Mexico City, with approval number 2014-3505-6 by the Ethics Committee. Informed consent signed by each participant, included their authorization for colposcopy, cervical scraping, biopsy sampling, HPV DNA detection and genotyping. The use of clinical data as well as sociodemographic and personal information of each participant was strictly confidential. Women were referred to colposcopy test, if a cervical anomalous result was diagnosed through gynecological examination or cytological test.

### 2.2. Patients

Three hundred and sixty-two women between 15–85 years old were eligible for the study. Data from women with HPV DNA positive test, cytologic diagnosis before colposcopy and complete clinical records were included in the study. Women with incomplete or missing clinical records, inadequate conditions for cervical scraping as menstruation, pregnancy or refusing to participate were not included in this study. The age of participants, age at first intercourse, total number of sexual partners, sexual intercourse without condoms, sexually transmitted diseases records (STD), age at first birth and smoking, were recollected from clinical reports.

### 2.3. Obtaining cervical samples and biopsy

During the gynecological examination, we collected two samples of cervical cells from all participants with cytobrush (both were immersed in two tubes with 2 ml of Preservcyt ®). One cytological samples was used for HPV DNA detection and genotyping and stored at −20°C until analysis. With the other cervical sample, we performed cytological smears and prepared for cytomorphological examination using Papanicolaou staining. In patients with suspected cervical high-grade lesion, a biopsy guided by colposcopy was performed (Colposcope with 50W LED illumination, Labomed Meditech). The biopsy was introduced into a vial with 4% buffered formalin and labelled with identification data. The colposcopy results were issued by a gynaecologist from the Department of Dysplasia Clinics. While cervical samples were diagnosed by the head of the Pathology Department of the same hospital. All samples were identified by consecutive number, name of participant (coded for confidentiality), grade of cervical lesion, and type of sample.

### 2.4. Diagnosis of cervical samples

Cytologic diagnoses formulated with the Bethesda System 2001 were negative for intraepithelial lesion or malignancy (NILM), atypical squamous cells of undetermined significance (ASCUS), low-grade squamous intraepithelial lesion (LSIL), high-grade squamous intraepithelial lesion (HSIL) and cervical cancer (CC) [17]. Histopathologic diagnoses according to the WHO criteria were: normal cervix (NC), cervical intraepithelial neoplasia grade 1 (CIN 1), cervical intraepithelial neoplasia grades 2, 3 and *in situ* (CIN 2/CIN 3 and *in situ*) and cervical cancer (CC) [18].

The cytological lesions were classified into two main categories:(1) ≤ LSIL, that included NILM, ASCUS, and LSIL, and (2) HSIL+, including HSIL and CC. Histological lesions were clustered in ≤ CIN1 (NC and CIN1) and CIN2+ (CIN2, CIN3/*in situ* and CC).

### 2.5. HPV DNA detection and genotyping

Genomic DNA was extracted from samples using a commercial kit (Wizard® Genomic DNA purification kit protocol, Promega) according to the manufacturer’s recommendations. To detect HR-HPV types 16, 18, 31, 33, 35, 39, 45, 51, 52, 56, 58, 59, 66 and 68, the purified DNA was then amplified by a strip probe assay [19] for the qualitative detection and identification of 32 genotypes of HPV by detecting specific sequences in the L1 region of the HPV genome grouped in HR-HPV, probable high risk (pHR: 26, 53, 70, 73, and 82) and low risk (LR: 6, 11, 40, 42, 43, 44, 54, 61, 62, 67, 81, 83, and 89). The genotyping is based on the principle of reverse hybridization. Part of the L1 region (SPF10 region) of HPV genome is amplified with specific primers, and the resulting biotinylated amplicons are denatured and hybridized with specific oligonucleotide probes. Additional primer is added for the amplification of the human HLA-DPB1 gene, to control the quality and extraction of the sample. All probes are immobilized as parallel bands in membrane strips. After hybridization and astringent washing, alkaline phosphatase conjugated with streptavidin is added, which binds to any previously formed biotinylated hybrid (INNO-LiPA HPV Genotyping *Extra II* Amp kit, Fujirebio) according to the manufacturer’s protocol.

### 2.6. Statistical analysis

To calculate the sample size we used single proportion test with a 95% confidence level [20]. Descriptive statistics were performed to summarize the sociodemographic characteristics of the selected population. In the same way, we estimate the overall HPV prevalence according to the cervical diagnosis. To determine the prevalence of HR-HPV types by cytologic diagnosis, we determined the specific proportion per samples positive to HR-HPV, in addition to cervical samples categories: HPV negative, low risk (LR-HPV), probable high risk (pHR-HPV), and high risk (HR-HPV) HPV infections, as reported previously [4]. The prevalence of HR-HPV in single infection and co-infection in cervical specimens was estimated by chi-squared test. We assessed the association among HR-HPV types according to cervical lesion (OR and _95%_CI chi-square test). Finally, to estimate the risk factors associated to the probability of infection by HR-HPV type we carried out bivariate and multivariate analysis adjusted (adOR, _95%_CI) by risk factors: age, age at first intercourse, total number of sexual partners, sexual intercourse without condom, sexually transmitted disease (STD) records, age at first birth, and smoking. All statistical tests were two-sided and considered significant at p<0.05. Statistical analysis was performed using commercial and open software (SPSS v25 and R, respectively).

## 3. Results

### 3.1. Sociodemographic and clinical characteristics of participants

Three hundred and sixty-two women met the eligibility criteria for the study. The median age was 40 years old (32-50). Their median age of beginning sexual life was 18 years (16-20) and 70% of them (253/362) have had two or more sexual partners. Of 362 participants 333 (92.0%) did not use a condom during their sexual relations and 61.0% (221/362) referred the antecedent of STD at some point in their life. The age of first birth in 60.5% (219/362) was before 19 years of age and 21.3% (77/362) reported smoking history (Table 1).

**Table 1.**
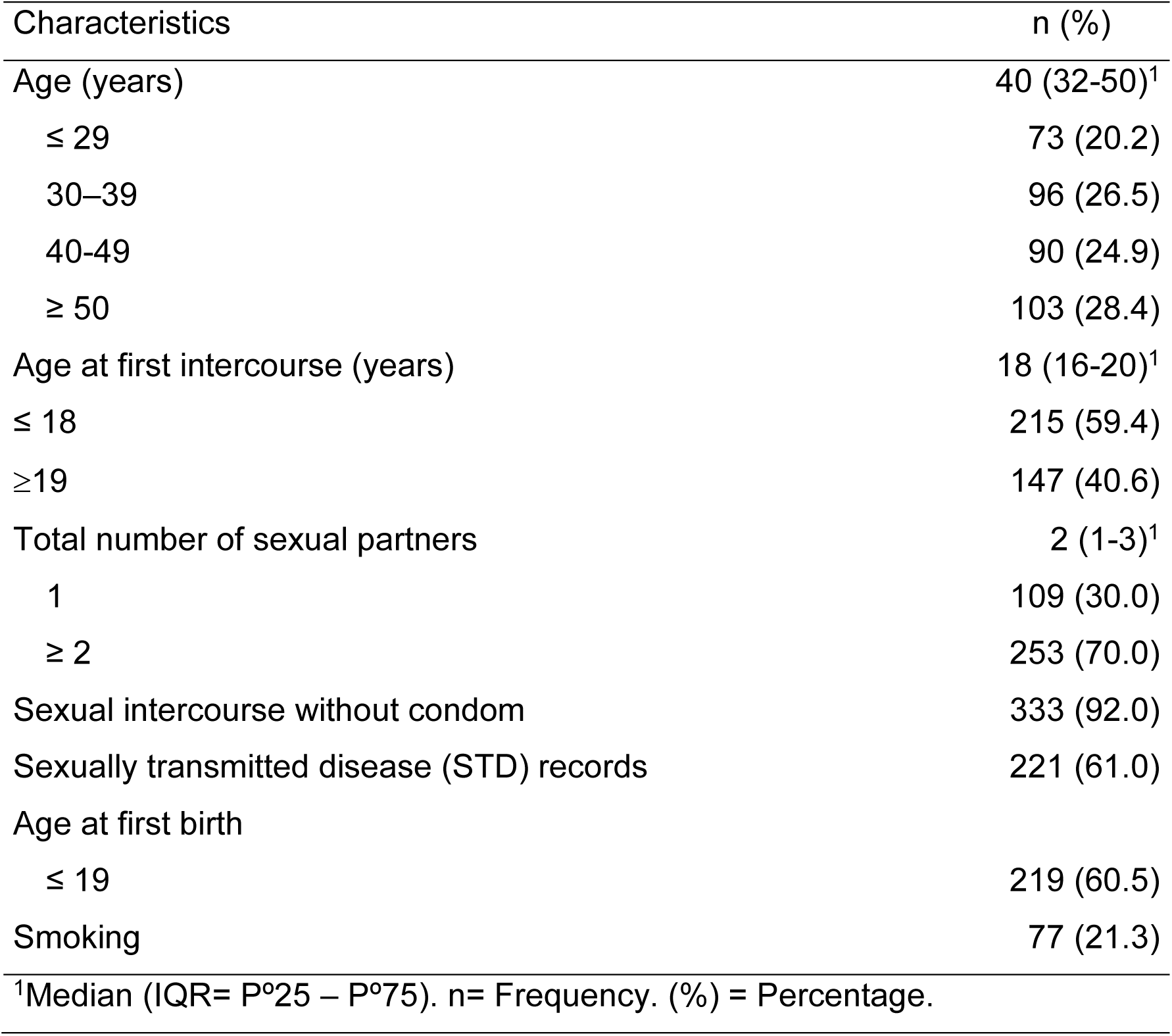
Sociodemographic and clinical characteristics of HPV positive Mexican women (n=362)

| Characteristics | n (%) |
| --- | --- |
| Age (years) | 40 (32-50)<sup>1</sup> |
| ≤ 29 | 73 (20.2) |
| 30–39 | 96 (26.5) |
| 40-49 | 90 (24.9) |
| ≥ 50 | 103 (28.4) |
| Age at first intercourse (years) | 18 (16-20)<sup>1</sup> |
| ≤ 18 | 215 (59.4) |
| ≥19 | 147 (40.6) |
| Total number of sexual partners | 2 (1-3)<sup>1</sup> |
| 1 | 109 (30.0) |
| ≥ 2 | 253 (70.0) |
| Sexual intercourse without condom | 333 (92.0) |
| Sexually transmitted disease (STD) records | 221 (61.0) |
| Age at first birth |  |
| ≤ 19 | 219 (60.5) |
| Smoking | 77 (21.3) |
| <sup>1</sup>Median (IQR= P<sup>0</sup>25 – P<sup>0</sup>75). n= Frequency. (%) = Percentage. |  |

### 3.2. HPV prevalence and HR-HPV types according to cervical diagnosis

In 362 cervical samples, 83.43% (302/362) were HR-HPV positive, 2.76% (10/362) were pHR-HPV positive, 4.97% (18/362) LR-HPV positive, and 8.84% (32/362) were HPV negative. In HR-HPV positive cervical samples, type 16 (46.7%) was the most prevalent, followed by types 66 (32.8%), 51 (31.8%), 52 (31.1%), 58 (20.2%), 59 (17.2%), and 56 (11.3%). Viral types with an overall prevalence lower than 10% were 18 (9.9%), 39 (8.3%), 45 (7.6%), 31 (5-0%), 68 (1.0%), 35 (1.0%), and 33 (0.7%) (Figure 1).

**Figure 1.**
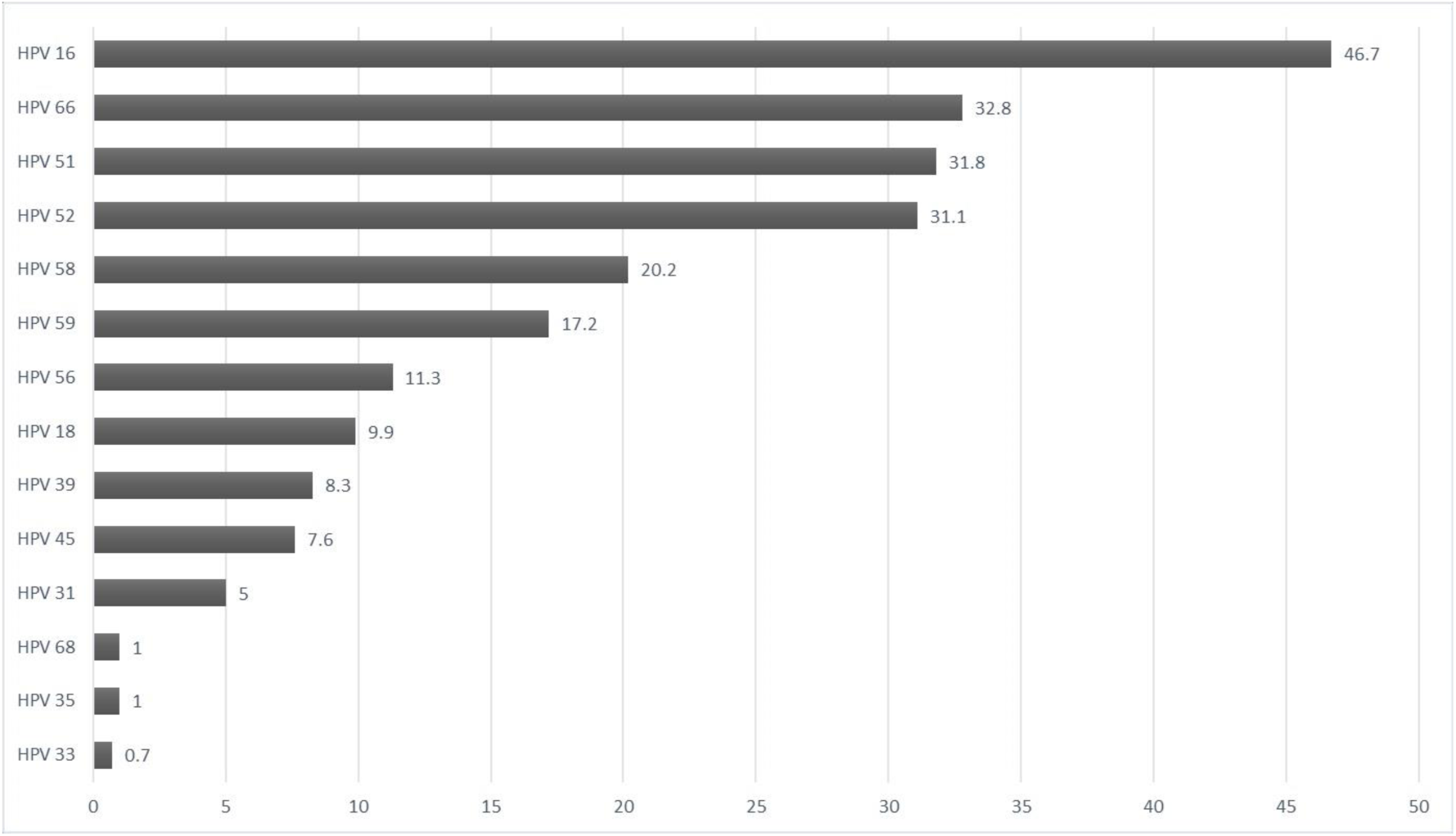
Prevalence of HPV types in positive cervical samples from Mexican women (n=302). The percentage values are shown

Cytological diagnosis included NILM (28, 9.3%), ASC-US (62, 20.5%), LSIL (105, 34.8%), HSIL (52, 17.2%), and CC (55, 18.20%). Two HPV types were the most prevalent analyzed by cytological diagnosis. HPV-16 had high prevalence in NILM, HSIL, and CC (78.6%, 50% and 58.2%, respectively), followed by HPV-66 in ASC-US and LSIL premalignant lesions (50% and 45.7%, respectively), and very low in CC (3.2%). In contrast, types 52 and 51 had approximately similar prevalence from NILM to HSIL, decreasing in CC. HPV-18 was present only in NILM and CC (25% and 14.5%, respectively). HPVs 45, 58 and 59 had heterogeneous prevalence among cytological categories. Finally, HPVs 31, 33, 35, 39, 56, and 68 had low prevalence and were absent in some cervical cytological categories. HPVs 51, 52, and 66 infections were predominantly observed among LSIL and ASC-US while HPV-16 was present in high percentages particularly in NILM and in CC (Figure 2).

**Figure 2.**
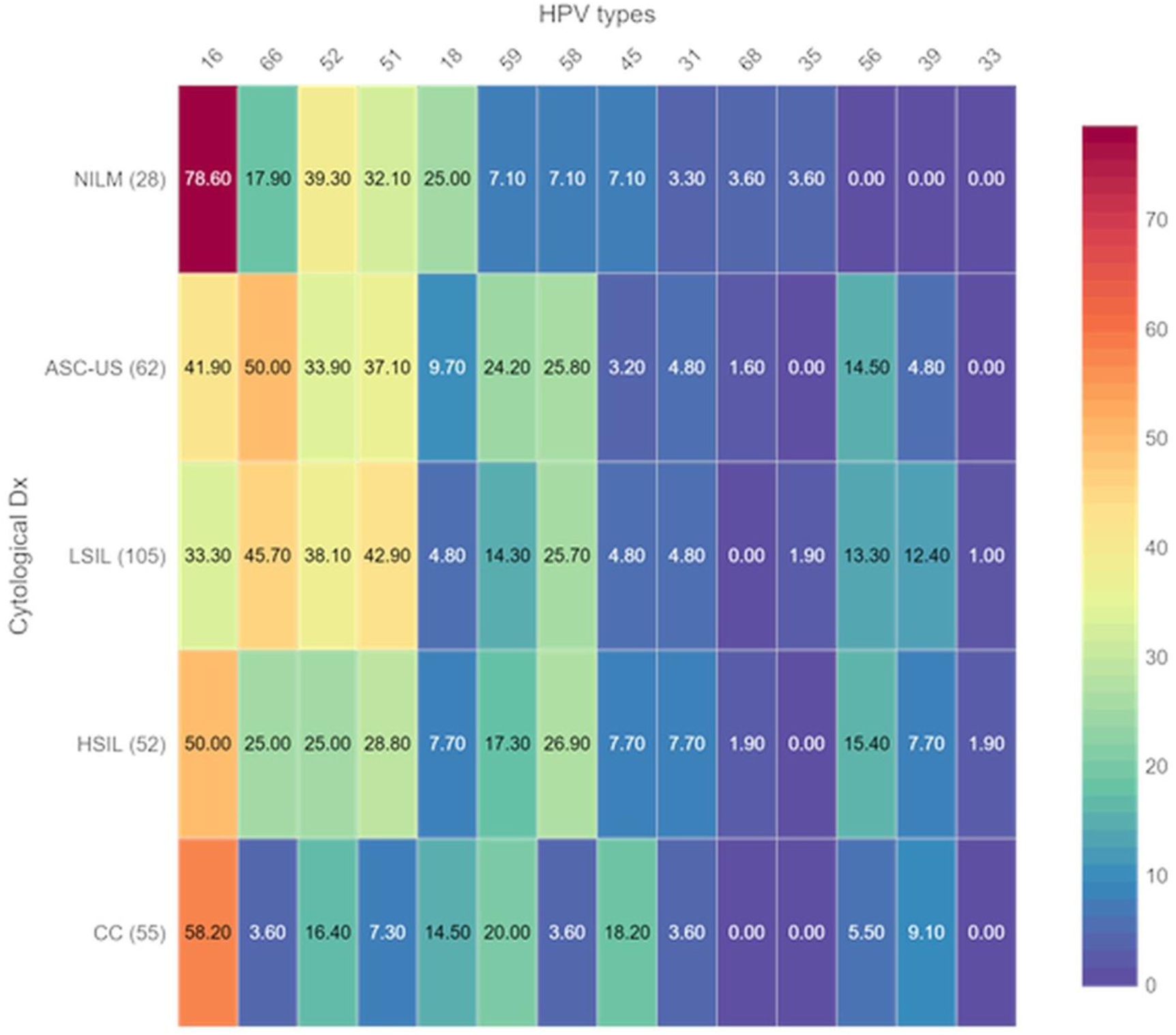
HR-HPV types according to cytologic diagnosis. The heatmap shows the prevalence in percentages between high-risk HPV types and diagnosis according to Bethesda classification. A color gradient is shown (right). Dx, diagnosis; NILM, negative for intraepithelial lesion or malignancy; ASC-US, atypical squamous cells of undetermined significance; LSIL, low-grade squamous intraepithelial lesion; HSIL, high-grade squamous intraepithelial lesion; CC, cervical cancer; human papillomavirus; HR-HPV, high risk HPV

### 3.3. HR-HPV prevalence in single and co-infection in cervical lesions

Of 302 cervical smears positive to HR-HPV, 38.4% (116/302) had single infection and 61.6% (186/302) had co-infection (≥ 2 HR-HPV types).

The most prevalent single infections involved HPV-16 (31.0%), followed by 52 (12.9%), 66 and 51 (10.3%, in both). Prevalence less than 10% was observed in HPV 39, 45 (7.8%, in both), 58 (5.2%), 18, 31, 56, and HPV 59 (3.4%, all), and 35 (0.9%). HPV 33 and 68 were absent. We performed a bivariate analysis (chi-square *test*, p<0.05), regardless of whether it was single or co-infection, and observed that specifically HPV-66 was associated to ASC-US (OR= 4.6, _95%_IC= 1.55-13.65, p< 0.05) and LSIL (OR= 3.87, _95%_ IC= 1.36-10.96). Regardless of the HR-HPV type, single infection was statistically significant in CC status (OR= 2.70, _95%_CI= 1.06-6.89, p< 0.05), and in HSIL+ (OR= 2.18, _95%_CI= 1.34-3.54, p< 0.05). The most frequent HR-HPV co-infections involved HPV 66-51 (28.49%), HPV 16-52 (25.80%), HPV 16-51 (23.65%), and HPV 16-66 (22.58%); other co-infections were below 20%. However, only two HR-HPV co-infections were significantly associated when assessing the probability of association (*X*^*2*^ test, p< 0.05): HPV 66-51 (OR= 3.41, _95%_CI= 1.86-6.26, p< 0.05) and HPV 52-58 (OR= 2.48, _95%_CI= 1.30-4.73, p< 0.05).

### 3.4. Associations among HR-HPVs according to cervical lesion

We found several significant associations (chi-square test, p< 0.05) between HR-HPV and cervical lesions. In LSIL, HPV 52-58 (OR= 4.04, _95%_CI= 1.4-11.64, p< 0.05) and HPV 66-51 (OR= 3.96, _95%_CI= 1.48-10.61, p<0.05) were significantly associated, while HPV 16-52 (OR= 7.87, _95%_CI= 1.35-45.83, p< 0.05) and HPV 66-51 (OR= 7.5, _95%_ CI= 1.49-37.65, p< 0.05) were in HSIL. When we analyzed HR-HPV risk infection by ≤ LSIL and HSIL+ categories, our results showed that in ≤ LSIL, HPV 66-51 and HPV 52-58 had a statistically significant association (OR= 2.818, _95%_CI= 1.388-5.720, p<0.05 and OR= 2.412, _95%_CI= 1.126-5.169, p< 0.05, respectively). Whereas in HSIL+, HPV 66-51 had a statistically significant association (OR= 5.22, _95%_CI= 1.411-19.31, p<0.05). Results according to the histopathological diagnosis of samples were similar to cytologic diagnosis (Data not shown).

### 3.5. Probability of HR-HPV infection and risk factors associated

In order to assess the HR-HPV risk infection by risk factors, we performed several bivariate and logistic regression models adjusted by the following risk factors (adOR, _95%_CI): age, age at first intercourse, total number of sexual partners, sexual intercourse without condom, sexually transmitted disease (STD) records, age at first birth, and smoking. We found that HPV-16 risk infection was statistically significant in women at 30-39 years old (adOR= 2.104, _95%_CI= 1.09-4.07, p< 0.05). Conversely, the risk of infection with types 66 and 52 was statistically significant in women with STDs records (adOR= 2.02, _95%_CI= 1.19-3.43, p< 0.05, adOR= 1.69, _95%_CI= 1.01-2.85, p< 0.05, respectively) (Figure 3).

**Figure 3.**
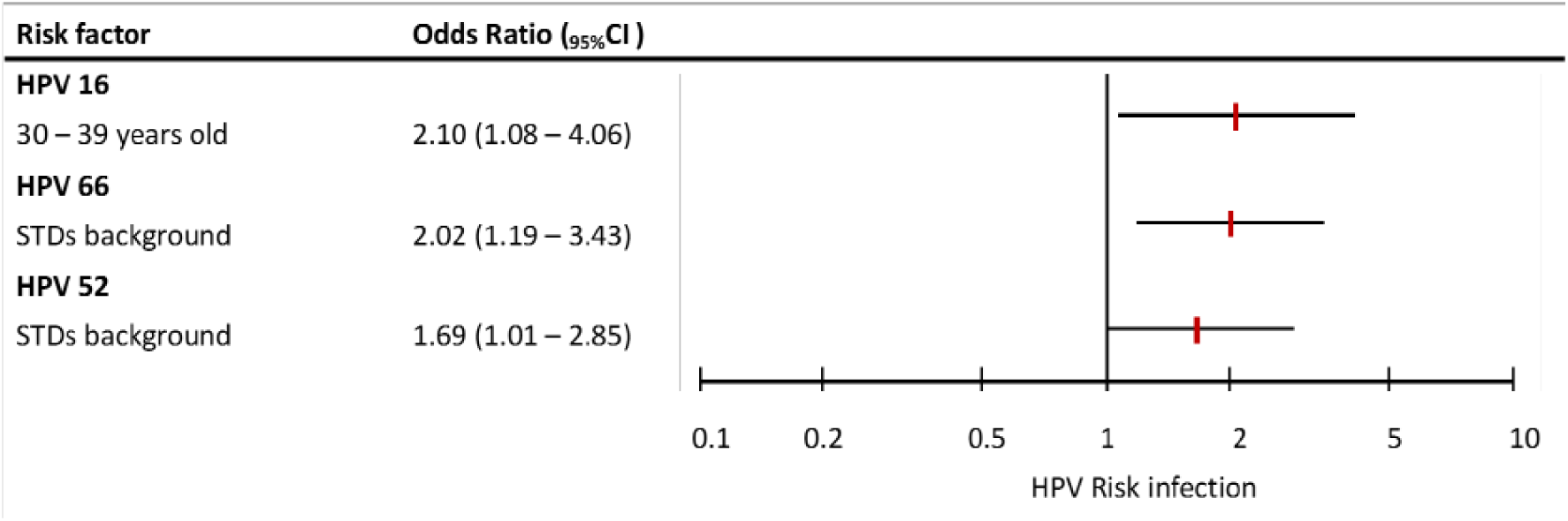
Risk factors associated with HPV infection in cervical samples of women referred to dysplasia clinics. Odds ratios adjusted (adOR) and 95% confidence interval (_95%_CI) for risk factors: age, age at first intercourse, total number of sexual partners, sexual intercourse without condom, sexually transmitted disease -STD-records, age at first birth, smoking. Only those HR-HPV and risk factors that had statistical significance for each logistic regression model are presented.

## 4. Discussion

In this cross-sectional study in Mexican women, we were able to identify 14 high-risk oncogenic HPV types in 83.43% of participants (302/364 abnormal cervical smears). Their sociodemographic characteristics were similar to those of previous studies [12,13,21]. However, HR-HPV overall prevalence (83.43%) was mildly higher in our study compared to previous reports such as those from Ortega Cervantes et al. (2016) [22] and Flores Miramontes et al. (2015) [23], with 82.0% and 71.24%, respectively. Our results showed that HPV-16 was the most frequent type (46.7%). Surprisingly, our study also showed a high prevalence of HPV-66 (32.8%), in contrast to other reports in which this type had a very low incidence (∼5%) [3]. According to Chouhy D. et al. (2013) [24] these results might be a consequence of the detection methods employed such as capturing hybrids that do not include HPV-66 or COBAS HPV test that detects it not specifically, but within a pool that includes other HR-HPVs [25,26]. Additionally, misidentifying this type by confusing it with type 56, such as with the linear array system [24]. In more recent reports using similar molecular techniques to detect HPV types, a higher prevalence of HPV-66 has been observed than in previous reports [12,13,21].

Also, other studies with HIV+ women and abnormal cytology reported high prevalence of HPV-66 (12.0% [27] and 32.3% [28]). Although, we did not have information concerning their immunological status of our participants, we had data of STD records and 60% of our study population had a history of STD. These set of diseases have been previously shown to be associated with HPV infection [29]; and in addition, we observed that STD was significantly associated with HPV types 52 and 66 infections.

HPV types such as 18, 51, 52, 58, and 59 showed similar prevalences to those previously reported in other studies in Mexican women [30,31]. Conversely, HPVs 31, 33, and 35, showed a lower prevalence than other studies from Mexico [32,33]. The differences observed in our study may be explained by geographical and regional HPV variations [34]. Besides, we should consider the effect of the migratory phenomenon that can cause an increase in the prevalence of other HPV genotypes [35].

In several studies HPV-66 has been detected in single infection, however other reports observed this viral type mainly in co-infection [21,34,36,37]. We found both situations in our study: co-infection and single infection in a 2:1 ratio. Regarding cervical lesion grade, HPV-66 presence was more important in ASC-US and LSIL (≤ LSIL) than in CC (HSIL+), which is in agreement with previous reports [37,38]. In co-infection events, HPV-66 had a statistically significant association with HPV-51 (p< 0.05), surprisingly not with HPV-16 (p> 0.05), even though is the most prevalent type in HPV infections.

Given these observations, in combination with previous studies where the contribution of HPV-66 to premalignant and malignant lesions of the uterine cervix was evaluated [39,40], HPV-66 infection might indicate a non-progressive to cervical cancer phenotype.

Regarding the risk factors analyzed, we agree that those associated with sexual activity were important for the acquisition of HR-HPV [5,41], in particular STD records. In this sense, despite sexually transmitted disease records were obtained through clinical files and the type of pathogen or infection was not specified, this limitation of our study might also be considered as a proposal for future research. A strength of our study was that the calculation of the sample size made it possible to obtain statistically reliable results regarding the real frequency of HPV-66 in Mexican women.

Finally, our evidence supports the notion that HPV-66 should not be considered as a high-risk type, since we found it predominantly in premalignant lesions and non-malignant samples. Additional studies are required to determine the real contribution of HPV-66 in cervical carcinogenesis and whether or not HPV-66 infection might indicate a non-progressive to cervical cancer phenotype.

## Conclusions

HPV-66 prevalence was unusually high in low-grade cervical lesions, predominantly in co-infection with HPV-51, and very low frequency in cervical cancer. This information should be taken into consideration for the interpretation of results obtained with methods that do not include or group type 66 with other HR-types. Finally, the presence of HPV-66 either in single or co-infections with other HR-HPV types, excluding 16 and 18 may, indicate lesions that will not progress to cancer. HPV-66 infection might be more frequent than previously thought, and specific genotyping may clarify these observations.

## Data Availability

All information about data VPH 66 is on SPSS/Base 20 08 19.sav

## Contribution of the authors

Equal contribution †= JG, PC.

Study conception and design: JG, PC, ME, MS, VV.

Acquisition of data: JG, PC, GR, GG, MV, MM, GC, ME, RR, SS, AS, PA, MS, VV.

Analysis of data: JG, PC, GG, GC, ME, RR, VV.

Interpretation of data: JG, PC, GG, MM, GC, ME, MS, VV.

Writing the article: JG, PC, GR, MV, ME, RR, SS, MS, VV.

Critical revision of the article: GR, GG, MV, MM, GC, ME, RR, SS, AS, PA, MS, VV.

Final approval of the article: JG, PC, GR, GG, MV, MM, GC, ME, RR, SS, AS, PA, MS, VV.

JG: Juárez González
PC: Paredes Cervantes
GR: Gordillo Rodríguez
GG: González Guzmán
MV: Moncayo Valencia
MM: Méndez Martínez
GC: García Carrancá
ME: Martínez Ezquerro
RR: Rivas Ruíz
SS: Sánchez Suárez
AS: Alvarez Sandoval
PA: Padilla Arrieta
MS: Martínez Salazar
VV: Vázquez Vega

## Acknowledgements

This project was supported by CONACyT Sectoral Funds (FOSISS) SALUD-2014-C01-234198. PROT/FIS/IMSS 1377. To the team of Cellular Pathology and Molecular Cancer Laboratory, UIMEO HO. CMN SXXI, IMSS.

This work was partially supported by a grant to AG-C from PAPIIT-UNAM (IN213016).

Manuscript Writing Training Team (CEMAI 2019-2) of CONACyT for their reviews and constructive criticism of this research paper.

